# Accounting for Twins and Other Multiple Births in Perinatal Studies Conducted Using Healthcare Administration Data

**DOI:** 10.1101/2024.01.23.24301685

**Authors:** Jeremy P Brown, Jennifer J Yland J, Paige L Williams, Krista F Huybrechts, Sonia Hernández-Díaz

## Abstract

The analysis of perinatal studies is complicated by twins and other multiple births even when they are not the exposure, outcome, or a confounder of interest. Common approaches to handling multiples in studies of infant outcomes include restriction to singletons, counting outcomes at the pregnancy-level (i.e., by counting if at least one twin experienced a binary outcome), or infant-level analysis including all infants and, typically, accounting for clustering of outcomes by using generalised estimating equations or mixed effects models. Several healthcare administration databases only support restriction to singletons or pregnancy-level approaches. For example, in MarketScan insurance claims data, diagnoses in twins are often assigned to a single infant identifier, thereby preventing ascertainment of infant-level outcomes among multiples. Different approaches correspond to different causal questions, produce different estimands, and often rely on different assumptions. We demonstrate the differences that can arise from these different approaches using Monte Carlo simulations, algebraic formulas, and an applied example. Furthermore, we provide guidance on the handling of multiples in perinatal studies when using healthcare administration data.

## Introduction

In the field of perinatal epidemiology, we inevitably encounter the question, “how should we handle twins?” The presence of multiples (i.e., twins and higher order multiples) can play many roles in research studies including as an exposure (e.g., what is the effect of multiple birth on postpartum hemorrhage?), an outcome (e.g., what is the effect of fertility treatment on multiple births?), or as a covariate. In the latter case, multiple gestation may be a confounder, mediator, collider, or effect modifier. In addition, multiples play an important role in statistical analysis even when they are independent of the causal pathway and not part of a causal question. Specifically, multiples create a clustered data structure because their outcomes are likely to be correlated. Thus, they require special consideration in both study design and statistical analysis.

For maternal treatments and infant outcomes, which are the focus of this study, common analytical approaches to handle multiples include 1) restriction to singletons, 2) counting binary outcomes at the pregnancy-level (by presence of at least one infant-level outcome), and 3) infant-level analyses including all infants and typically accounting for the effect of clustering of outcomes within multiples by using generalised estimating equations (GEE) or mixed effects models.^1,2^ Estimates can differ across these approaches.

Due to data recording and reporting practices, perinatal studies using healthcare administration data are often limited to either restriction to singletons or pregnancy-level analyses. For example, in the MarketScan Commercial Claims and Encounters (MarketScan CCAE) United States (US) insurance claims data, diagnoses among twins are often recorded to a single infant enrolment identifier, especially for siblings of the same sex, or may be recorded in the maternal records, especially for in utero diagnoses (see eAppendix for a detailed description of multiples data in MarketScan CCAE). In this circumstance, it is often only possible to determine that at least one infant experienced an outcome.

While the role of multiple births has been previously considered, particularly in trials as a source of clustering,^3–10^ there has been limited attention paid to the difference between estimands and assumptions needed when restricting to singletons or counting outcomes at the pregnancy-level. Our aim is to illustrate differences between approaches, through Monte Carlo simulations, algebraic formulas, and an empirical applied example, and to provide guidance for perinatal studies using healthcare administration data.

## Methods

### Approaches to handle multiple births

We compare three approaches to handle multiple births in perinatal studies with an infant outcome: restriction to singletons, counting outcomes at the pregnancy-level, and infant-level analysis including all infants.

One approach that can universally be applied and is commonly implemented in perinatal studies is restriction to singletons. This approach eliminates any issue of clustering of outcomes and problems with assigning outcomes to an infant. However, when multiple gestation is a post-treatment mediator (Figure 1a and 1b), collider (Figure 1c), or interacts with treatment then the estimated effect can differ from the average infant-level effect including all infants because we condition on multiple births through restriction. If pre-conception treatment is associated with increased risk of multiple gestation (e.g. fertility treatments), and there are uncontrolled common causes of multiple gestation and outcome, multiple gestation will be a collider and conditioning on it can introduce selection bias even when the effect of treatment on the outcome is null (Figure 1c).^11^ Alternatively, if multiple gestation is a mediator (Figure 1a and 1b), restricting to singleton births (a proxy for single gestation), will largely eliminate the effect of treatment mediated by multiple gestation, which may be of clinical and public health interest as part of the total effect of treatment on the outcome.^12,13^ For a post-conception treatment, multiple gestation may be a confounder (Figure 1d - e.g., multiples may confound the effect of caesarean section on NICU admission), which could be controlled by restricting to single gestation pregnancies, stratifying by number of fetuses, or otherwise adjusting for multiple gestation.

**Figure 1:**
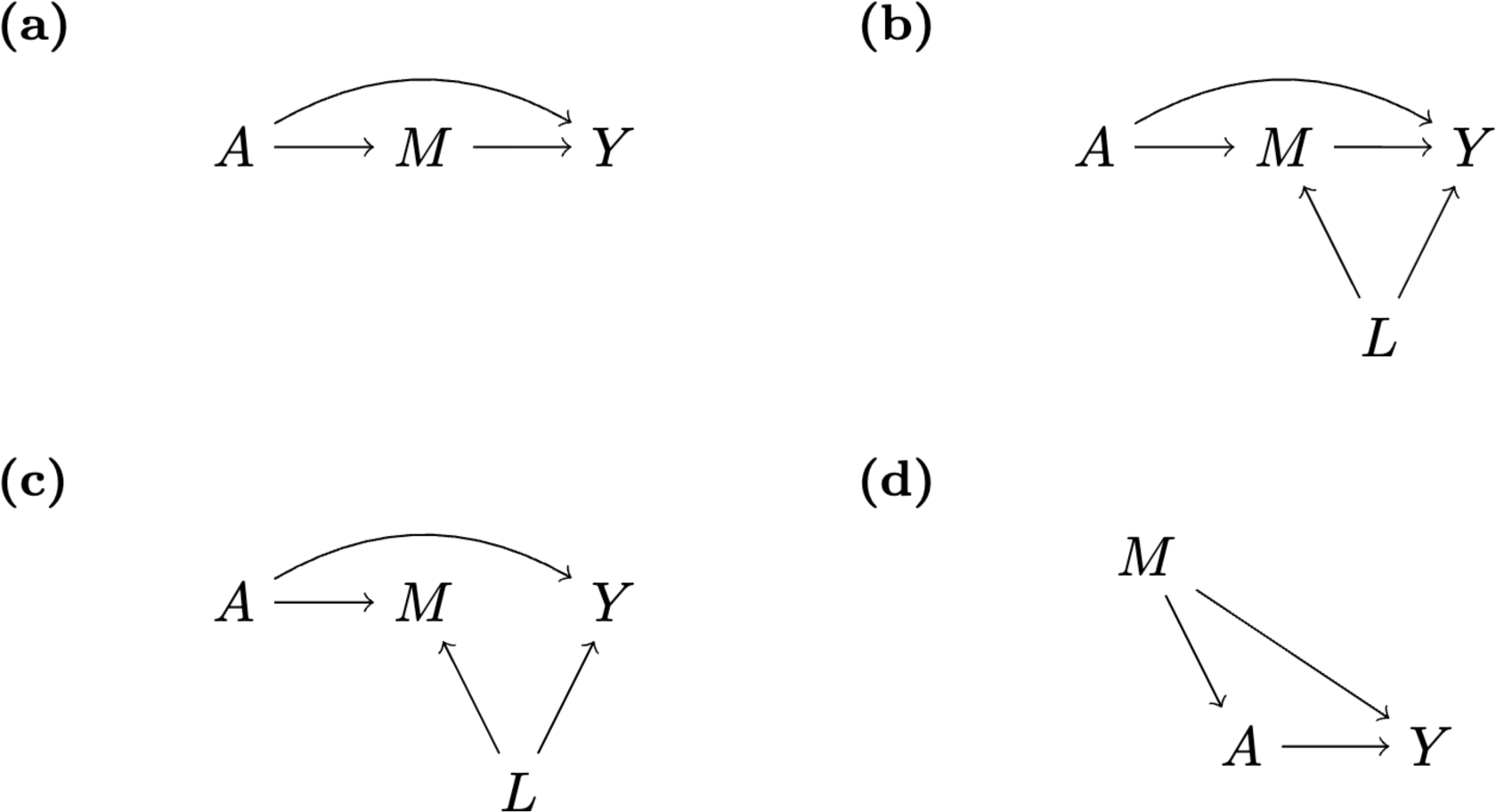
Directed acyclic graphs with treatment A, outcome Y, multiple gestation M, and confounder L illustrating the potential role of multiple gestation as a) a mediator, b) a mediator with mediator-outcome confounding (e.g., by maternal age), c) a collider, or d) a confounder

A second approach that can be applied for binary outcomes in the absence of infant-level outcome data is to count outcomes at the pregnancy-level by recording whether at least one infant experienced an outcome. In this approach, the risk of an outcome for a twin pregnancy will typically be higher (up to 2x) than the infant-level risk among all infants, purely because there are more opportunities for the outcome to occur. The magnitude of increased risk depends on the degree to which the outcomes of the twins within each pregnancy are correlated (see eAppendix for algebraic comparison). This introduces an edge in the directed acyclic graph between multiple gestation and the outcome. Thus, given a treatment-multiples association, multiple gestation may become a mediator^14^, if treatment is pre-conception, or a confounder, if treatment is post-conception. Either situation may result in an association between treatment and outcome at the pregnancy-level, even if there is no association at the infant-level.

We focus in this article on analyses restricted to pregnancies ending in live birth. These analyses, which condition on conceiving and surviving until birth, introduce some assumptions for causal inference, in particular absence of selection bias.^11,15^ Analyses including women who do not conceive or give birth are inherently at the maternal or pregnancy-level (e.g., among prospective mothers planning pregnancy what is the risk with one intervention versus another of both conceiving, having a live birth, and having at least one infant with a malformation?). For preconception treatments and live-birth outcomes, this risk will be affected by the effect of treatment on conception, multiples, and pregnancy loss, in addition to to any direct effect on the outcome.

Infant-level analyses including all infants are commonly applied in studies using prospective data collection but are more challenging to apply when using healthcare administration data. When infant-level analyses are feasible, one difficulty is that multiple births introduce clustering of outcomes. This clustering violates independence assumptions that are typically made and can therefore lead to inaccurate variance estimates. GEE or mixed effect models can be used to obtain more efficient estimators and more accurate quantification of uncertainty in clustered data.^1,2,16^ Both approaches have been used in pregnancy studies.^3–6^ It is important to note that these two approaches often estimate different effects, with mixed effect models, unlike GEE, typically estimating effects conditional on cluster-level random effects.^17^ However, approaches to marginalise over these cluster-level random effects are available.^18^ Furthermore, with GEE, different assumptions regarding the assumed correlation structure lead to different estimands if cluster-size is informative for the outcome.^19^

Perinatal studies are atypical for clustered studies in that treatment and covariates, which are both measured at the cluster-level (i.e., the pregnancy), can subsequently affect cluster-size. The distribution of maternal covariates at the infant-level when including multiples can differ from the distribution of covariates at the pregnancy-level if covariates are associated with multiple pregnancy (see eFigure 1 for illustrative tree diagram). When analysing preconception treatments and calculating marginal treatment effects, we will typically want to achieve balance in the covariate distribution at the pregnancy-level (e.g. by inverse probability of treatment weighting) as we would in a randomised controlled trial.^20^ Balance at the pregnancy-level will not necessarily imply balance on the covariate distribution at the infant-level when treatment affects multiples even in randomised trials, given that treatment may affect the covariate distribution in infants through an effect on twinning.

### Monte Carlo simulations

We used Monte Carlo simulations to compare differences between these three approaches (i.e., restriction to singletons, pregnancy-level approach, infant level-approach) under a range of data-generating mechanisms. Binary random variables were simulated from Bernoulli distributions for a binary pregnancy-level treatment *A*, a binary infant outcome *Y*, and a binary pregnancy-level indicator of multiple gestation *M.* Multiple gestation was simulated to be a post-treatment covariate and potential mediator as in our applied example (Figure 1b). For simplicity of exposition, we consider that all multiple gestations are twin pregnancies and ignore pregnancy losses. In practice, multiple gestation will be a non-zero count, but the frequency of higher order pregnancies (>2) is rare.^21^ Treatment and multiple gestation were simulated at the pregnancy-level. Following this, each row of data with multiple gestation was duplicated and infant outcomes were simulated. For the second twin, the outcome was simulated using the method of Qaqish et al. to generate a variable with identical mean, but such that the intraclass correlation among twins between the first infant, Y_1_, and the second infant, Y_2_, is equal to α.^22^

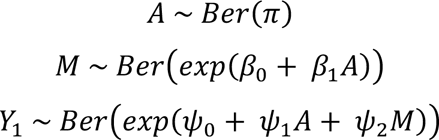

Parameter values were chosen to reflect values observed in perinatal studies. For the different scenarios we fixed the number of pregnancies at 10,000, treatment prevalence (π) at 50%, and the prevalence of outcome among unexposed singletons (exp (*ψ*_0_)) at 3%, and the prevalence of multiples among unexposed pregnancies (exp(*β*_0_)) at 2%, and varied factorially the risk ratio between treatment and multiples (exp(*β*_1_)), the conditional risk ratio between treatment and outcome (exp (*ψ*_1_)), the conditional risk ratio between multiple gestation and outcome (exp (*ψ*_2_)), and the intraclass correlation coefficient (ICC) α (see Table 1).

**Table 1:** Parameter values chosen in Monte Carlo simulations.

| Parameter | Description | Values |
| --- | --- | --- |
| $\pi$ | Prevalence of treatment | 0.5 |
| $\exp(\beta_0)$ | Prevalence of multiple gestation among untreated pregnancies | 0.02 |
| $\exp(\beta_1)$ | Risk ratio between treatment and multiples | 1 to 10 in increments of 0.5 |
| $\exp(\psi_0)$ | Prevalence of outcome amongst untreated singletons | 0.03 |
| $\exp(\psi_1)$ | Risk ratio between treatment and outcome conditional on multiple gestation | 1,2 |
| $\exp(\psi_2)$ | Risk ratio between multiples and outcome conditional on treatment | 1,2 |
| $\alpha$ | Intraclass correlation coefficient | 0,0.5,1 |

For restriction to singletons and pregnancy-level analyses, Poisson models were fitted to estimate risk ratios with Huber-White heteroskedasticity-robust standard errors.^23,24^ For the infant-level analysis, we applied Poisson GEE with an independence working correlation structure, which should more robustly estimate the variance of effect estimates in the presence of clustering.^1^ For each scenario, we generated 1,000 simulated datasets to which we applied analyses. We compare, between scenarios, the exponentiated mean of the log risk ratio.

### Applied example

We empirically illustrated the differences between methods using an example of fertility treatment and infant outcomes. Specifically, we evaluated the risk of major congenital malformations and neonatal intensive care unit (NICU) admission, comparing pregnancies/infants conceived via assisted reproductive technology (ART), relative to two comparator groups: 1) pregnancies/infants conceived via intrauterine insemination (IUI), and 2) pregnancies/infants conceived without IUI or ART. The latter example was selected for didactic purposes since the intervention (fertility treatment) is strongly associated with multiples. However, this comparison is more prone to residual confounding. We have previously compared ART to IUI, in a target trial emulation framework, to address clinical questions.^25,26^

We conducted the study using MarketScan CCAE, a US database of insurance claims for private health insurance beneficiaries.^27^ In this database, insurance claims for twins of the same sex are commonly recorded to the same enrolment identifier. Within MarketScan we identified a cohort of pregnancies linked to liveborn infants delivered by women between the ages of 12 and 55 years from 2011 to 2021. We used validated algorithms to identify pregnancies, estimate gestational age, and link mother-infant pairs.^28–32^ For each pregnancy, we required maternal enrolment and prescription eligibility from at least 90 days before the last menstrual period (LMP) until 30 days after delivery. The treatment of interest, ART, and comparator treatment, IUI, were defined based on the presence of procedure codes recorded between LMP - 30 days and LMP + 30 days. Covariates included were potential confounders ascertained from maternal claims, which included maternal age, clinical conditions, lifestyle variables, and dispensed medications (see eAppendix for details and code lists).

The outcomes of interest were major congenital malformation and neonatal intensive care unit (NICU) admission. Malformations occur in about 3% of infants, with some evidence for increased risk among multiples in particular for monozygotic twins.^36–41^ NICU is a relatively common outcome (10-15% of infants are admitted to the NICU^33^), highly correlated within pregnancies^34^, and is affected by multiple gestation given increased frequency of preterm births among twin pregnancies.^35^ Within-pregnancy correlation is anticipated to be lower for malformations than for NICU admission, with concordance among twins with one or more non-chromosomal malformations of 11.4% observed in a large European study of 5.4 million births.^42^

Major congenital malformation, an outcome previously validated in US claims data,^43^ was defined as a structural abnormality of surgical, medical, or cosmetic importance. To identify malformations, we required infant enrolment from birth to at least 90 days after delivery, unless the infant died sooner. We identified International Classification of Diseases (ICD) 9/10 diagnosis and procedure codes indicating a birth defect or a corrective surgical procedure in infant claims during the 90 days after birth or in maternal claims around the time of delivery.^43^ We required at least one code in the infant record to identify a malformation. NICU admission was identified using CPT (Current Procedural Terminology) codes in the infant records within 30 days after delivery.

Multiple births were identified by either two linked infants or the presence of more than one multiple gestation code after first trimester in the absence of a mixed birth code (i.e., combined stillbirth and live birth). Given that in MarketScan CCAE claims are commonly recorded among twins to a single infant identifier, we were limited in our analyses, and compared restriction to singletons to counting at the pregnancy-level. Poisson models with Huber-White standard errors were fitted to estimate conditional risk ratios adjusting for covariates.^24^ Age was modelled with linear and quadratic terms. Among singletons, and at the pregnancy-level, outcomes can be assumed to be independent, and therefore particular statistical methods to account for clustering were unneccesary.

A subset of twins were linked to two infant identifiers. As a secondary analysis, to approximate an infant-level effect we excluded pregnancies classified as multiple births but with only one linked infant identifier and calculated an inverse probability of selection weighted treatment effect. Probability of selection was estimated at the pregnancy-level using a logistic regression model including treatment and all covariates. Inverse probability of selection weighted Poisson GEE was applied with an independence working correlation structure to estimate conditional risk ratios adjusted for covariates.

As a sensitivity analysis, rather than restrict to singleton births, we restricted to pregnancies with no record of multiple gestation, excluding pregnancies with a multiple gestation code in the first trimester only and mixed births. This is not anticipated to substantially impact findings, given that only a small fraction of singleton births are expected to be from multiple gestation pregnancies (i.e., due to loss of one twin).

Data management was performed using SAS Enterprise Guide 8.3 with statistical analyses and Monte Carlo simulations conducted using R 4.2.0. Code used for statistical analyses and simulations is available at https://github.com/CausalInference/CAUSALab_Papers.

## Results

### Monte Carlo simulations

In simulations, we compared the effect of restriction to singletons and counting outcomes at the pregnancy-level relative to infant-level analyses including all infants.

For restriction to singletons, estimates differed from the infant-level analyses including all infants when there was mediation by multiple gestation (Figure 2 D-F, J-L). In these scenarios, restriction to singletons eliminated the effect mediated by multiple gestation. The difference between the treatment effects depended on the strength of mediation, as determined by the treatment-multiples association, and was minor when the association between treatment and multiple gestation was small.

**Figure 2:**
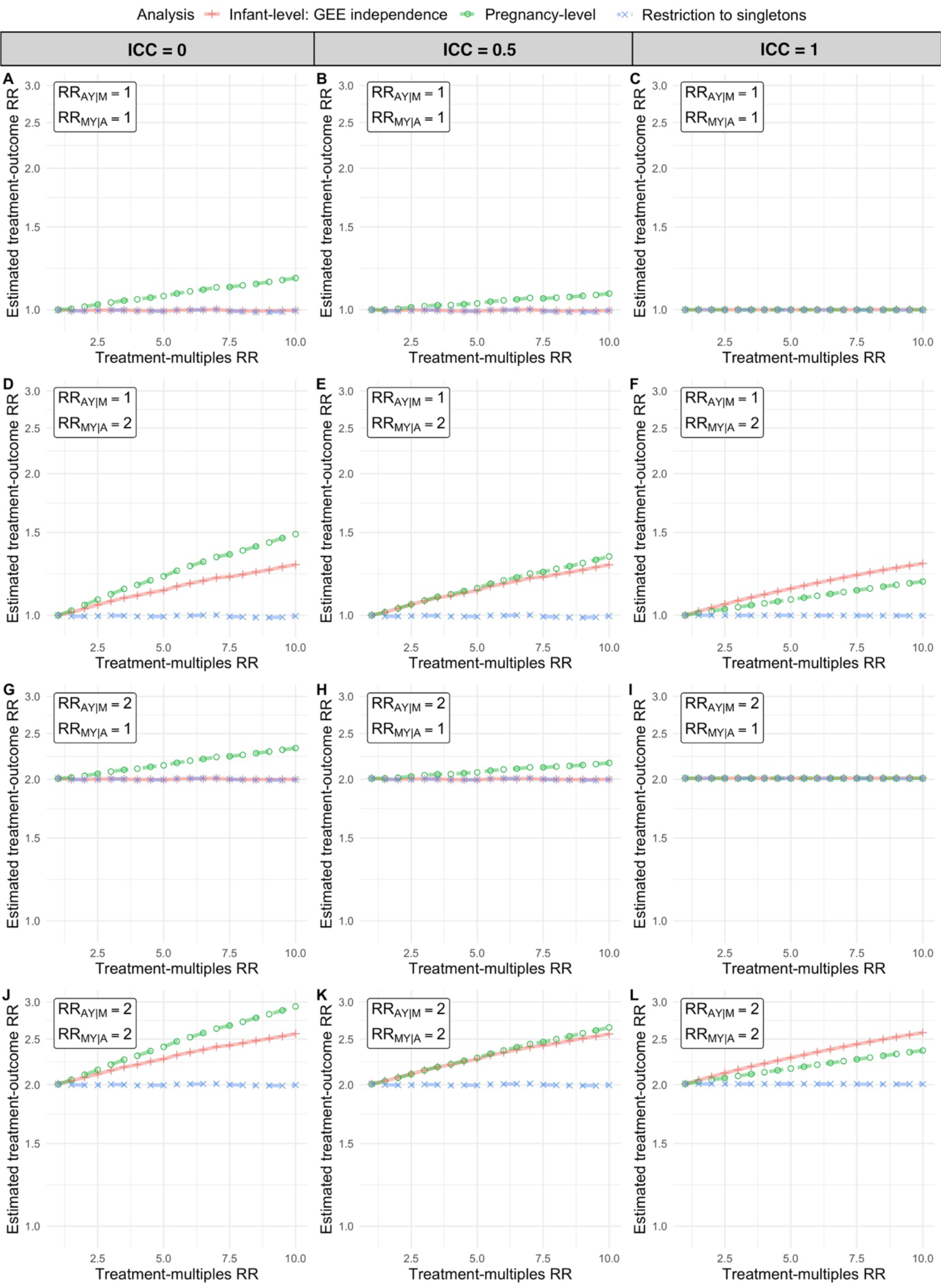
Estimated risk ratios in Monte Carlo simulations with treatment-outcome conditional risk ratio (*RR_AY|M_*) of 1 (A-F) or 2 (G-L), multiple gestation-outcome conditional risk ratio (*RR_AY|M_*) of 1 (A-C, G-I) or 2 (D-F, J-L), and intraclass correlation coefficient (ICC) of 0 (A, D, G, J), 0.5 (B, E, H, K) or 1 (C, F, I, L)

For pregnancy-level analyses, estimates differed from infant-level analyses including all infants depending on the association between treatment and multiple gestation and the correlation of the outcome within a pregnancy. When treatment was associated with increased multiple gestation, the pregnancy-level risk ratio exceeded the infant-level estimate when the outcomes were independent or moderately correlated (Figure 2 A, B, D, E, G, H, J, K), and was lower than the infant-level risk ratio when outcomes were highly correlated and multiple gestation was associated with the outcome conditional on treatment (Figure 2 F & L). All approaches led to the same estimates when the ICC was 1 and there was no mediation by multiple gestation (Figure 2 C & I). As is apparent from an algebraic analysis (see eAppendix), the difference between approaches in risk of an outcome in a treatment group will depend on the prevalence of multiples, the prevalence of the outcome, and the correlation of the outcome within a multiple birth. The results of simulations match results from algebraic formulas (see eAppendix for spreadsheet implementing formulas).

### Applied example

We selected a cohort of 903,273 pregnancies linked to live-born infants, of which 25,634 (2.8%) were to women treated with ART, 10,600 (1.2.%) to women treated with IUI, and 867,039 (96.0%) to women not treated with IUI or ART (Table 2).

**Table 2:** Characteristics of pregnancies to women treated with ART, IUI or neither fertility treatment.

| **Characteristic** | **Non-use**<br>N = 867,039 | **ART**<br>N = 25,634 | **IUI**<br>N = 10,600<sup>1</sup> |
| --- | --- | --- | --- |
| Age (years) |  |  |  |
| ≤19 | 1,187 (0.1) | 0 (0.0) | 0 (0.0) |
| 20-29 | 257,770 (29.7) | 1,989 (7.8) | 1,430 (13.5) |
| 30-39 | 568,546 (65.6) | 18,601 (72.6) | 8,375 (79.0) |
| 40-49 | 39,536 (4.6) | 5,044 (19.7) | 795 (7.5) |
| Multiple birth | 18,333 (2.1) | 4,031 (15.7) | 1,198 (11.3) |
| Any multiple gestation | 22,868 (2.6) | 4,916 (19.2) | 1,415 (13.3) |
| Linked to two infants IDs | 6,280 (0.7) | 1,725 (6.7) | 535 (5.0) |
| Tobacco use | 15,450 (1.8) | 364 (1.4) | 130 (1.2) |
| Alcohol abuse or dependence | 1,359 (0.2) | 48 (0.2) | 16 (0.2) |
| Drug abuse or dependence | 4,595 (0.5) | 95 (0.4) | 40 (0.4) |
| Obesity<sup>a</sup> | 97,644 (11.3) | 3,457 (13.5) | 1,607 (15.2) |
| Definite or suspected teratogen | 55,725 (6.4) | 2,135 (8.3) | 632 (6.0) |
| Antidepressants | 71,426 (8.2) | 1,869 (7.3) | 788 (7.4) |
| Anticonvulsants | 10,958 (1.3) | 259 (1.0) | 124 (1.2) |
| Opioids | 75,769 (8.7) | 5,940 (23.2) | 1,000 (9.4) |
| Codeine | 20,637 (2.4) | 2,062 (8.0) | 269 (2.5) |
| Antibiotics | 175,620 (20.3) | 17,640 (68.8) | 3,284 (31.0) |
| Epilepsy | 3,716 (0.4) | 121 (0.5) | 46 (0.4) |
| Hypertension | 44,382 (5.1) | 1,523 (5.9) | 539 (5.1) |
| Diabetes | 34,351 (4.0) | 4,429 (17.3) | 1,567 (14.8) |

a) Claims-based marker of obesity, which is expected to underestimate true obesity prevalence.
Definitions: ART, assisted reproductive technology; ID, identifier; IUI, intrauterine insemination.

Women treated with ART or IUI were older than women not treated with IUI or ART (92% aged 30+ for ART, 87% for IUI, and 70% for neither treatment) and had higher prevalence of diabetes (17.3% for ART, 14.8% for IUI, and 4.0% for neither). Both ART and IUI were associated with an increase in multiple births, with a frequency of 15.7% among ART pregnancies and 11.3% among IUI pregnancies, relative to 2.1% among pregnancies not treated with either therapy.

Of the 4,031 ART pregnancies with a multiple birth, less than half (N=1,725, 42.3%) were linked to two infant identifiers. Given the recording of claims in Marketscan CCAE from same sex twins to a single infant identifier, among these 1,725 pregnancies 1,720 (99.7%) of the twin pairs had divergent sex (i.e. one male and one female). Among pregnancies with at least one outcome, the concordance rate was 12.1% (23 of 190 pregnancies) for malformation and 71.9% (586 of 815 pregnancies) for NICU. Furthermore, the ICC was 0.16 (0.12-0.21) for malformations and 0.72 (0.70-0.75) for NICU. The frequency per infant identifier among multiples linked to two identifiers was 6.2% for malformation and 40.6% for NICU admission (Table 3); the corresponding frequencies among singletons were 6.1% and 13.2%. The frequency of at least one outcome among these multiple birth pregnancies linked to two identifiers was 11.0% (190 of 1725 pregnancies) for malformation and 47.2% (815/1,725) for NICU. For multiple birth pregnancies linked to a single infant identifier, the frequency per identifier was 11.4% for malformation and 44.0% for NICU.

**Table 3:** Outcome incidence per infant identifier among singletons, multiple births linked to a single infant identifier, and multiple births linked to two infant identifiers.

| **Outcome** | **Multiple birth**<br>**(1 linked infant),**<br>**N = 2,307<sup>a</sup>** | **Multiple birth**<br>**(2 linked infants),**<br>**N = 3,450<sup>b</sup>** | **Singleton,**<br>**N = 21,603** |
| --- | --- | --- | --- |
| Any malformation | 263 (11.4%) | 213 (6.2%) | 1,320 (6.1%) |
| Non-chromosomal malformation | 249 (10.8%) | 197 (5.7%) | 1,278 (5.9%) |
| Central nervous system | 35 (1.5%) | 20 (0.6%) | 76 (0.4%) |
| Cardiac | 76 (3.3%) | 49 (1.4%) | 307 (1.4%) |
| Gastrointestinal | 29 (1.3%) | 28 (0.8%) | 117 (0.5%) |
| Genital | 33 (1.4%) | 32 (0.9%) | 248 (1.1%) |
| Urinary | 38 (1.6%) | 34 (1.0%) | 185 (0.9%) |
| Muscular | 37 (1.6%) | 35 (1.0%) | 205 (0.9%) |
| Limb | 27 (1.2%) | 16 (0.5%) | 151 (0.7%) |
| NICU admission | 1,015 (44.0%) | 1,401 (40.6%) | 2,857 (13.2%) |

a) From 2,307 pregnancies
b) From 1,725 pregnancies
Definitions: NICU, neonatal intensive care unit.

The association between ART and outcomes relative to non-use of ART/IUI differed by analytical approach with an adjusted risk ratio for malformation of 1.17 (95% CI 1.10-1.23) when restricting to singletons, 1.29 (95% CI 1.23-1.35) when counting at pregnancy-level, and 1.19 (95% CI 1.12-1.26) when estimating an infant-level effect including multiples using inverse probability of selection weights (Figure 3 – see eFigure 2 for crude estimates). For NICU admission, the adjusted risk ratios were 1.27 (95% CI 1.22-1.32) when restricting to singletons, 1.57 (95% CI 1.52-1.61) when counting at the pregnancy-level, and 1.69 (95% CI 1.63-1.76) when estimating an infant-level effect including multiples.

**Figure 3:**
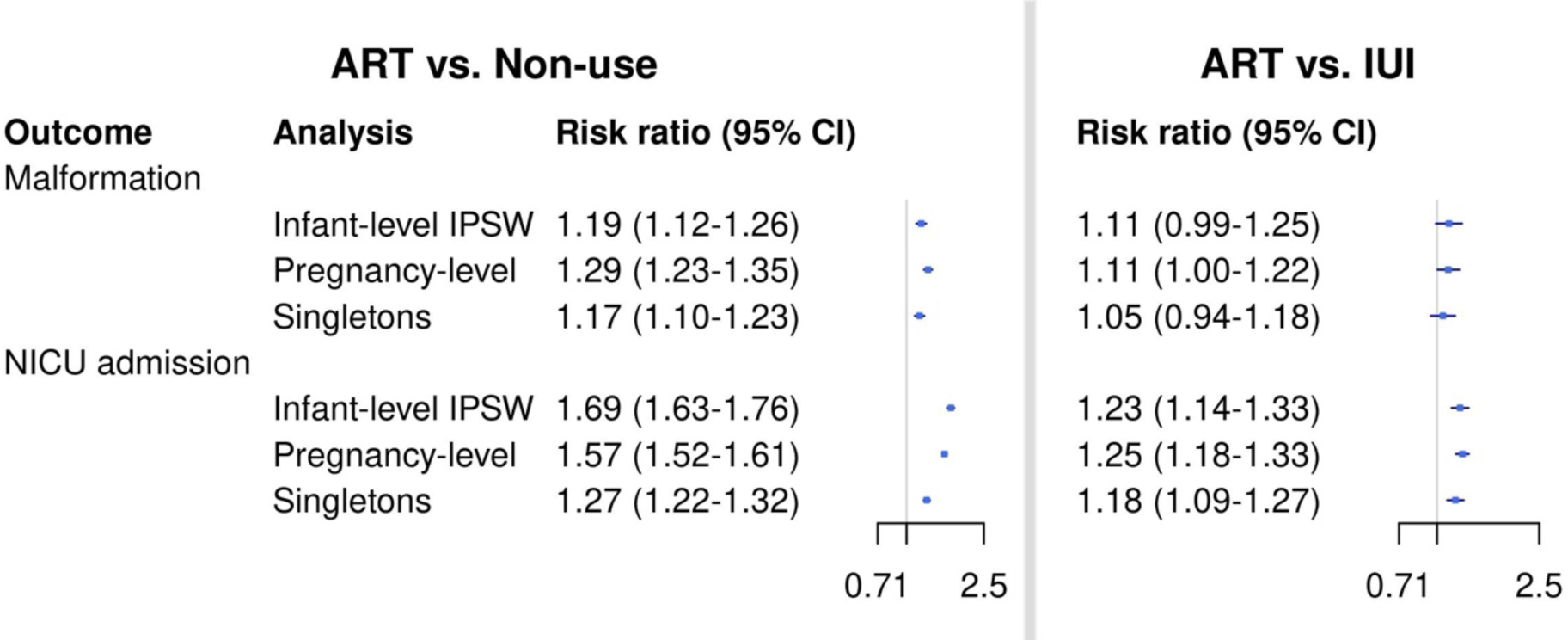
Adjusted risk ratios between assisted reproductive technology (ART) and both malformation and neonatal intensive care unit (NICU) admission relative to intrauterine insemination (IUI) and ART/IUI non-use

In comparison, there were smaller differences between analytical approaches when ART was compared with IUI, a treatment similarly associated with increased multiples (Figure 3). The adjusted risk ratio for malformation was 1.05 (95% CI 0.94-1.18) when restricting to singletons, 1.11 (95% CI 1.00-1.22) when counting at pregnancy-level, and 1.11 (95% CI 0.99-1.25) when calculating an infant-level effect including multiples. Similarly, for NICU the estimates were similar with a risk ratio of 1.18 (95% CI 1.09-1.27) when restricting to singletons, 1.25 (95% CI 1.18-1.33) when counting at the pregnancy-level, and 1.23 (95% CI 1.14-1.33) when estimating the infant-level effect. In a sensitivity analysis, restriction to pregnancies without any record of multiple gestation differed little relative to restriction to singleton births (eTable 1).

## Discussion

When designing perinatal studies with infant outcomes using healthcare administration data, it is valuable to not only ask what question is of interest and which estimand this corresponds to, but also which analyses are feasible given the data. With answers to these questions, an informed choice of analysis can be made. In simulations and our applied example, estimates from different approaches were similar when multiples were rare and either not associated or weakly associated with treatment, but differed substantively when treatment and multiples were strongly associated.

When multiple gestation is a mediator, restriction to singletons eliminates some or all of the effect of treatment on outcome mediated by multiple gestation, as was apparent in the Monte Carlo simulations and in the applied example. Residual confounding due to infertility may explain the remaining association in singletons. When multiple gestation is a mediator, if the question of interest is the etiological effect of treatment separate from multiples, restriction to singletons can approximate this effect subject to mediation analysis assumptions.^12^ However, if the total effect is of interest, infant-level analyses including all infants is advisable if feasible.

For binary outcomes, counting outcomes at the pregnancy-level by presence of at least one outcome addresses a different question: What is the risk for a mother of having one or more infants with the outcome? As is apparent from the algebraic formulas, simulations, and the applied example, estimates from this approach can differ from an infant-level analysis, particularly when treatment is associated with multiples. In the applied example comparing ART to non-use, for malformations, an outcome with weak intraclass correlation, the adjusted risk ratio was elevated in pregnancy-level analysis relative to the association in infant-level analysis including multiples (1.29 [95% CI 1.23-1.35] vs. 1.19 [95% CI 1.12-1.126]). For NICU admission, an outcome with strong intraclass correlation, the adjusted risk ratio was lower relative to the infant-level association including multiples (1.57 [95% CI 1.52-1.61] vs. 1.69 [95% CI 1.63-1.76]). These empirical findings accord with the results of the Monte Carlo simulations. Associations relative to IUI were more similar between approaches, reflecting the weaker association between treatment and multiples. However, it is important to note that even if relative risk is minimally affected, absolute risks in both treatment groups will typically be higher in pregnancy-level analyses.

Estimates of infant-level risk including multiples will commonly be an estimand of interest, but analyses to estimate these are not always feasible when using healthcare administration data. The recording of outcomes from multiple infants to a single infant identifier in Marketscan CCAE was apparent from examining the relation between multiplicity and outcome in the ART treatment group.

As a secondary analysis in the applied example, we estimated an infant-level effect including multiples by excluding multiple births linked to a single infant identifier and reweighting multiple births linked to two infant identifiers using inverse probability of selection weights. The validity of this approach depends on selection not being informative for the outcome given covariates, and that there is no cross-registration of outcomes between infants with two identifiers. The similarity between the concordance rate among twins for malformations linked to two infant identifiers of 12.1% in this study relative to the concordance rate of 11.4% in a large European registry study provides some reassurance on the validity of this infant-level data.^42^ Furthermore for NICU admission, the ICC of 0.72 is within the range of estimates from twelve randomized trials of 0.56-0.86 (median 0.79).^34^ One reason why censoring may be informative is that same sex multiples are more frequently linked to a single infant identifier in MarketScan data and will include all monozygotic twins, who may have a different risk of outcome than dizygotic twins. Additional assumptions needed are that triplets or other higher order multiples are rare, which is typically the case, and that multiple births are correctly classified.

When infant-level analyses are feasible, the question of what estimand is of interest remains. GEE with an independence correlation structure will estimate an average infant-level effect over all infants. While GEE with an independence correlation structure allows us to ask the question of what the average risk is per infant among all infants of women exposed to one treatment versus another, we can use inverse cluster-weighted GEE to address a slightly different question:^19^ What is the typical risk for a typical pregnancy? In this approach each pregnancy, rather than each infant, is weighted equally when calculating average risk. This estimand may be of interest, particularly for understanding and communicating the expected risk for a prospective mother.

## Conclusion

In summary, our analyses highlight the importance of understanding both the study question and the role of multiples in a study. In healthcare administration data, we are further limited in choice of analysis by the data. When multiples are rare in both treatment groups, as may be the case in many studies, different approaches to handle multiples are likely to produce similar estimates, but when multiples are more common or are associated with treatment, such as for fertility treatments, choice of approach is important and can produce differences between estimates.

## Data Availability

MarketScan CCAE is a commercial claims insurance database available by commercial license from Merative.

## Ethics Approval

This study was deemed exempt from review by the Harvard T.H. Chan School of Public Health Institutional Review Board.

## References

1. Liang KY, Zeger SL. Longitudinal data analysis using generalized linear models. Biometrika. 1986;73(1):13–22. doi:10.1093/biomet/73.1.13

2. Laird NM, Ware JH. Random-Effects Models for Longitudinal Data Nan M. Laird; James H. Ware Biometrics, Vol. 38, No. 4. (Dec., 1982), pp. 963-974. Biometrics. 1982;38(4):963–974.

3. Gates S, Brocklehurst P. How should randomised trials including multiple pregnancies be analysed? BJOG: Int J Obstet Gynaecol. 2004;111(3):213–219. doi:10.1111/j.1471-0528.2004.00059.x

4. Ananth CV, Platt RW, Savitz DA. Regression Models for Clustered Binary Responses: Implications of Ignoring the Intracluster Correlation in an Analysis of Perinatal Mortality in Twin Gestations. Ann Epidemiology. 2005;15(4):293–301. doi:10.1016/j.annepidem.2004.08.007

5. Ananth CV. Epidemiologic Approaches for Studying Recurrent Pregnancy Outcomes: Challenges and Implications for Research. Semin Perinatol. 2007;31(3):196–201. doi:10.1053/j.semperi.2007.03.008

6. Marston L, Peacock JL, Yu K, et al. Comparing methods of analysing datasets with small clusters: case studies using four paediatric datasets. Paediatr Périnat Epidemiology. 2009;23(4):380–392. doi:10.1111/j.1365-3016.2009.01046.x

7. Yelland LN, Scurrah KJ, Ferreira P, et al. Conducting Clinical Trials in Twin Populations: A Review of Design, Analysis, Recruitment and Ethical Issues for Twin-Only Trials. Twin Res Hum Genet. 2021;24(6):359–364. doi:10.1017/thg.2021.52

8. Yelland LN, Sullivan TR, Pavlou M, Seaman SR. Analysis of trials with informative birth size. Paediatr Périnat Epidemiology. 2015;29(6):567–575. doi:10.1111/ppe.12228

9. Yelland LN, Sullivan TR, Makrides M. Accounting for multiple births in randomised trials: a systematic review. Arch Dis Child - Fetal Neonatal Ed. 2015;100(2):F116. doi:10.1136/archdischild-2014-306239

10. Yelland LN, Salter AB, Ryan P, Makrides M. Analysis of binary outcomes from randomised trials including multiple births: when should clustering be taken into account? Paediatr Périnat Epidemiology. 2011;25(3):283–297. doi:10.1111/j.1365-3016.2011.01196.x

11. Hernán MA, Hernández-Díaz S, Robins JM. A Structural Approach to Selection Bias. Epidemiology. 2004;15(5):615–625. doi:10.1097/01.ede.0000135174.63482.43

12. VanderWeele TJ. Mediation Analysis: A Practitioner’s Guide. Annu Rev Publ Health. 2015;37(1):1–16. doi:10.1146/annurev-publhealth-032315-021402

13. Oberg AS, VanderWeele TJ, Almqvist C, Hernandez-Diaz S. Pregnancy complications following fertility treatment—disentangling the role of multiple gestation. Int J Epidemiology. 2018;47(4):1333–1342. doi:10.1093/ije/dyy103

14. Wen SW, Miao Q, Taljaard M, et al. Associations of Assisted Reproductive Technology and Twin Pregnancy With Risk of Congenital Heart Defects. JAMA Pediatr. 2020;174(5):446–454. doi:10.1001/jamapediatrics.2019.6096

15. Chiu YH, Stensrud MJ, Dahabreh IJ, et al. The Effect of Prenatal Treatments on Offspring Events in the Presence of Competing Events: An Application to a Randomized Trial of Fertility Therapies. Epidemiology. 2020;31(5):636–643. doi:10.1097/ede.0000000000001222

16. Ziegler A, Kastner C, Blettner M. The Generalised Estimating Equations: An Annotated Bibliography. Biom J. 1998;40(2):115–139. doi:10.1002/(sici)1521-4036(199806)40:2<115::aid-bimj115>3.0.co;2-6

17. Hubbard AE, Ahern J, Fleischer NL, et al. To GEE or Not to GEE. Epidemiology. 2010;21(4):467–474. doi:10.1097/ede.0b013e3181caeb90

18. Heagerty PJ, Zeger SL. Marginalized multilevel models and likelihood inference (with comments and a rejoinder by the authors). Stat Sci. 2000;15(1):1–26. doi:10.1214/ss/1009212671

19. Williamson JM, Datta S, Satten GA. Marginal Analyses of Clustered Data When Cluster Size Is Informative. Biometrics. 2003;59(1):36–42. doi:10.1111/1541-0420.00005

20. Robins JM, Hernán MÁ, Brumback B. Marginal Structural Models and Causal Inference in Epidemiology. Epidemiology. 2000;11(5):550–560. doi:10.1097/00001648-200009000-00011

21. Centers for Disease Control and Prevention, Cent N. National Vital Statistics System, Natality on CDC WONDER Online Database.

22. Qaqish BF. A family of multivariate binary distributions for simulating correlated binary variables with specified marginal means and correlations. Biometrika. 2003;90(2):455–463. doi:10.1093/biomet/90.2.455

23. Huber PJ. The Behavior of Maximum Likelihood Estimates Under Nonstandard Conditions. Berkeley Symp on Math Statist and Prob. Published online 1967.

24. White H. A Heteroskedasticity-Consistent Covariance Matrix Estimator and a Direct Test for Heteroskedasticity. Econometrica. 1980;48(4):817. doi:10.2307/1912934

25. Chiu YH, Yland JJ, Rinaudo P, et al. Effectiveness and safety of intrauterine insemination vs. assisted reproductive technology: emulating a target trial using an observational database of administrative claims. Fertil Steril. 2022;117(5):981–991. doi:10.1016/j.fertnstert.2022.02.003

26. Hernán MA, Wang W, Leaf DE. Target Trial Emulation. Jama. 2022;328(24):2446–2447. doi:10.1001/jama.2022.21383

27. Butler AM, Nickel KB, Overman RA, Brookhart MA. Databases for Pharmacoepidemiological Research. Springer Ser Epidemiology Public Heal. Published online 2021:243–251. doi:10.1007/978-3-030-51455-6_20

28. MacDonald SC, Cohen JM, Panchaud A, McElrath TF, Huybrechts KF, Hernández-Díaz S. Identifying pregnancies in insurance claims data: Methods and application to retinoid teratogenic surveillance. Pharmacoepidem Drug Safe. 2019;28(9):1211–1221. doi:10.1002/pds.4794

29. Zhu Y, Thai TN, Hernandez-Diaz S, et al. Development and Validation of Algorithms to Estimate Live Birth Gestational Age in Medicaid Analytic eXtract Data. Epidemiology. 2023;34(1):69–79. doi:10.1097/ede.0000000000001559

30. Chui YH, Huybrechts K, Zhu Y, Bateman B, Logan R, Hernandez-Diaz S. Internal validation of gestational age estimation algorithms in healthcare databases using pregnancies conceived with fertility procedures. In Press. American Journal of Epidemiology.

31. Margulis AV, Setoguchi S, Mittleman MA, Glynn RJ, Dormuth CR, Hernández-Díaz S. Algorithms to estimate the beginning of pregnancy in administrative databases. Pharmacoepidem Drug Safe. 2013;22(1):16–24. doi:10.1002/pds.3284

32. Li Q, Andrade SE, Cooper WO, et al. Validation of an algorithm to estimate gestational age in electronic health plan databases. Pharmacoepidemiol Drug Saf. 2013;22(5):524–532. doi:10.1002/pds.3407

33. Braun D, Braun E, Chiu V, et al. Trends in Neonatal Intensive Care Unit Utilization in a Large Integrated Health Care System. JAMA Netw Open. 2020;3(6):e205239. doi:10.1001/jamanetworkopen.2020.5239

34. Yelland L, Schuit E, Zamora J, et al. Correlation between neonatal outcomes of twins depends on the outcome: secondary analysis of twelve randomised controlled trials. BJOG: Int J Obstet Gynaecol. 2018;125(11):1406–1413. doi:10.1111/1471-0528.15292

35. Gardner M, Goldenberg R, Cliver S, Tucker J, Nelson K, Copper R. The origin and outcome of preterm twin pregnancies. Obstet Gynecol. 1995;85(4):553–557. doi:10.1016/0029-7844(94)00455-m

36. Centers for Disease Control and Prevention. Update on overall prevalence of major birth defects--Atlanta, Georgia, 1978-2005. MMWR Morbidity and mortality weekly report. 2008;57(1):1–5.

37. Herskind AM, Pedersen DA, Christensen K. Increased Prevalence of Congenital Heart Defects in Monozygotic and Dizygotic Twins. Circulation. 2013;128(11):1182–1188. doi:10.1161/circulationaha.113.002453

38. Hardin J, Carmichael SL, Selvin S, Lammer EJ, Shaw GM. Increased prevalence of cardiovascular defects among 56,709 California twin pairs. Am J Méd Genet Part A. 2009;149A(5):877-886. doi:10.1002/ajmg.a.32745

39. Hall JG. Twinning. Lancet. 2003;362(9385):735–743. doi:10.1016/s0140-6736(03)14237-7

40. J. DM, M. MV, J. WK, et al. Reproductive Technologies and the Risk of Birth Defects. N Engl J Med. 2012;366(19):1803–1813. doi:10.1056/nejmoa1008095

41. Zhu JL, Basso O, Obel C, Bille C, Olsen J. Infertility, infertility treatment, and congenital malformations: Danish national birth cohort. BMJ. 2006;333(7570):679. doi:10.1136/bmj.38919.495718.ae

42. Boyle B, McConkey R, Garne E, et al. Trends in the prevalence, risk and pregnancy outcome of multiple births with congenital anomaly: a registry-based study in 14 European countries 1984–2007. BJOG: Int J Obstet Gynaecol. 2013;120(6):707–716. doi:10.1111/1471-0528.12146

43. Cooper WO, Hernandez-Diaz S, Gideon P, et al. Positive predictive value of computerized records for major congenital malformations. Pharmacoepidem Drug Safe. 2008;17(5):455–460. doi:10.1002/pds.1534

